# J-difference GABA-edited MRS reveals altered cerebello-thalamo-cortical metabolism in patients with hepatic encephalopathy

**DOI:** 10.1101/2022.09.28.22280460

**Authors:** Helge J. Zöllner, Thomas A. Thiel, Nur-Deniz Füllenbach, Markus S. Jördens, Sinyeob Ahn, Dieter Häussinger, Markus Butz, Hans-Jörg Wittsack, Alfons Schnitzler, Georg Oeltzschner

## Abstract

Hepatic encephalopathy (HE) is a common neurological manifestation of liver cirrhosis. Clinical symptoms range from subtle attention deficits and motor disturbance to stupor and hepatic coma in the most severe cases. HE pathophysiology is characterized by an increase of ammonia in the brain due to impaired clearance in the cirrhotic liver. This results in disturbed glutamate-glutamine homeostasis as ammonia is increasingly metabolized by glutamine synthetase. Ammonia accumulation furthermore causes increased oxidative stress and disrupts neurotransmitter balance, including the GABAergic and glutamatergic systems. Clinical symptoms in the motor domain suggest that the cerebello-thalamo-cortical system plays a key role in HE. The aim of this study is to investigate metabolic abnormalities in the cerebello-thalamo-cortical system of HE patients using GABA-edited MRS. The study also investigates links between metabolite levels, disease severity, critical flicker frequency (CFF), motor performance scores, and blood ammonia levels.

GABA-edited MRS was performed in 35 participants (16 controls, 19 patients (3 minimal HE, 16 HE)) on a clinical 3T MRI system. MRS voxels were placed in the right cerebellum, left thalamus, and left motor cortex. GABA+ levels were estimated from the GABA-edited difference spectra using Gaussian fitting with the Gannet software. Levels of other metabolites of interest (glutamine, glutamate, myo-inositol, glutathione, total choline, total NAA, and total creatine) were assessed using linear-combination modeling in LCModel. Creatine- and water-referenced levels were reported to minimize biases of both reference standards. Group differences in metabolite levels and associations with clinical metrics were tested. Modeling uncertainty estimates of metabolite levels (Cramer-Rao Lower Bounds) were included as statistical weighting factors.

GABA+ levels were significantly increased in the cerebellum of patients with HE. GABA+ levels in the motor cortex were significantly decreased in HE patients, and correlated with the CFF (r = 0.73; p < .05) and motor performance scores (r = −0.65; p < .05). Well-established HE-typical metabolite patterns (increased glutamine, decreased myo-inositol and total choline) were confirmed in all three regions. These alterations were closely linked to clinical metrics. Increased glutathione levels were found in the thalamus and motor cortex. Explorative analysis indicated increased aspartate levels in all three regions and decreased scyllo-inositol levels in the motor cortex.

In summary, our findings provide further evidence for alterations in the GABAergic system in the cerebellum and motor cortex in HE. These changes were accompanied by characteristic patterns of osmolytes and oxidative stress markers in the cerebello-thalamo-cortical system. These metabolic disturbances are a likely contributor to HE motor symptoms in HE.

**Graphical Abstract:** 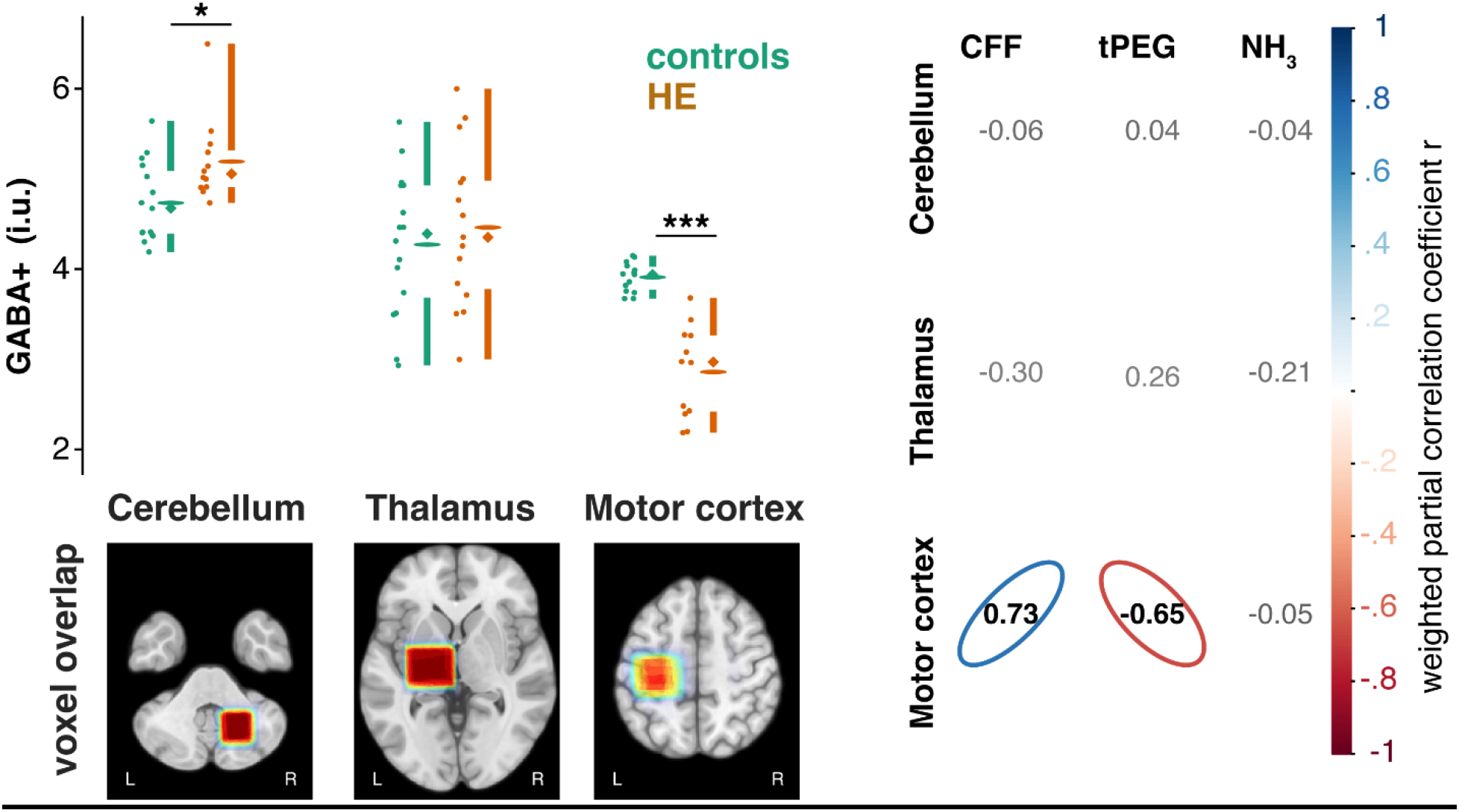

In patients with hepatic encephalopathy, GABA+ levels in the cerebello-thalamo-cortical loop are significantly increased in the cerebellum and significantly decreased in the motor cortex. GABA+ levels in the motor cortex strongly correlate with critical flicker frequency (CFF) and motor performance score (pegboard test tPEG), but not blood ammonia levels (NH_3_).

**Highlights:**

- Motor deficits in HE may originate from the cerebello-thalamo-cortical system
- Altered GABAergic neurotransmission plays a critical role in the pathophysiology of HE
- J-difference GABA-edited MRS can be used to study in vivo GABA+ levels
- Cerebellar and motor cortical GABA+ levels were significantly altered in HE
- GABA+ levels in the motor cortex strongly correlated with clinical metrics

## Introduction

Hepatic encephalopathy (HE) is a systemic neurological manifestation of liver cirrhosis indicating poor prognosis (Bustamante et al., 1999; Häussinger et al., 2022, 2021; Stewart et al., 2007). HE patients commonly suffer from altered cognitive performance, sleep disturbances, and impaired motor function and behavior. These clinical symptoms manifest with greatly varying severity during disease progression, starting with subtle attentional deficits, cognitive deterioration, disorientation, and disturbed motor control. The progression may conclude with somnolence, stupor, and in the most severe cases, hepatic coma (Butterworth, 2000; Felipo, 2013; Ferenci et al., 2002; Prakash and Mullen, 2010).

The pathophysiology is not fully understood but is considered to be multi-factorial (Cichoz-Lach and Michalak, 2013; Felipo, 2013; Häussinger et al., 2021). Cerebral hyperammonemia triggered by the blood-brain-barrier passage of blood ammonia is considered to be a key contributor. Ammonia accumulation results in numerous systemic responses, including neuroinflammation, low-grade edema, and oxidative stress (Cudalbu and Taylor-Robinson, 2019; Detry et al., 2006; Häussinger et al., 2021; Häussinger and Schliess, 2008; Norenberg et al., 2005; Pierzchala et al., 2022; Simicic et al., 2022). The main site of ammonia detoxification are the astrocytes in which glutamine synthetase scavenges ammonia in the glutamate-glutamine cycle. This process leads to strongly elevated glutamine (Gln) levels that can be detected with magnetic resonance spectroscopy (MRS) (Norenberg et al., 2005). It is hypothesized that Gln accumulation disrupts osmotic balance in the astrocytes, ultimately resulting in low-grade edema (Cudalbu and Taylor-Robinson, 2019; Detry et al., 2006; Häussinger et al., 2021; Häussinger and Schliess, 2008) that might even be detectable with MR imaging (Oeltzschner et al., 2016; Shah et al., 2008). Gln accumulation is further thought to cause compensatory depletion of the osmotic regulator myoinositol (mI) (Häussinger et al., 2021). Increased Gln and reduced mI levels reflecting the astroglial response to hyperammonemia is a well-established MRS pattern in HE (Häussinger et al., 1994; Kreis et al., 1991; Pathania et al., 2020; Zeng et al., 2020).

Altered GABAergic inhibition is considered to play a crucial role in the pathophysiology of HE (Häussinger et al., 2021; Sørensen et al., 2022). They were initially assumed to be a ‘brain-wide’ phenomenon resulting in a uniform GABAergic tone across the brain (Schafer and Anthony Jones, 1982). However, recent studies of hyperammonemia animal models reported region-specific changes to GABAergic neurotransmission (Cauli et al., 2009a). This was further substantiated by in-vivo human studies using GABA-edited MRS, suggesting region-specific changes of in-vivo GABA levels in HE, including the visual cortex and the striatum (Oeltzschner et al., 2015; Zupan et al., 2019). However, in vivo measurement of this low-concentration neurotransmitter remains technically challenging at clinical field strength (<= 3T), limiting its widespread application in HE. J-difference GABA-edited MRS allows improved resolution of the GABA signal by applying a frequency-selective pulse during the acquisition that reveals the 3-ppm GABA resonance, which is usually covered by much larger resonances (Choi et al., 2021; Mescher et al., 1998; Rothman et al., 1993). This technique has been successfully applied to elucidate the role of GABA in numerous neuroscientific and clinical studies (Bollmann et al., 2015; Foerster et al., 2013; Gao et al., 2013; Mostofsky et al., 2015; Muthukumaraswamy et al., 2009; Porges et al., 2017; Puts et al., 2011).

Oxidative stress is considered another important factor in HE pathophysiology (Häussinger et al., 2021; Simicic et al., 2022). In the healthy brain, reactive oxygen species (ROS) are formed during normal oxygen metabolism. As ROS can cause severe damage to cells, several complex systems regulate redox homeostasis to maintain ROS levels. The natural response to oxidative stress is the oxidization of glutathione (GSH), the most abundant antioxidant in the human brain. Afterwards, reduced GSH is regenerated from the oxidized form to maintain the oxidative potential of the cell and protect cellular integrity (Hilgier et al., 2010). This leads to increased total GSH levels, as observed in cell culture experiments, animal studies, and in vivo MRS studies at 3T (Murthy et al., 2000; Oeltzschner et al., 2016; Węgrzynowicz et al., 2007). Specifically, elevated total GSH levels were found in the cerebellum and cortex of rodents after acute liver failure (Sathyasaikumar et al., 2007) and in the motor cortex of HE patients (Oeltzschner et al., 2016). The cerebello-thalamo-cortical system is generally considered to be the major brain network involved in motor learning and fine motor control (Evarts and Thach, 1969; Holmes, 1939). It has been suggested that cerebellar disturbances propagate through the network resulting in worsening goal-directed movement (Holmes, 1939). Animal models of hyperammonemia and acute liver failure indicate disturbances in the GABAergic and glutamatergic neurotransmitter systems in the cerebello-thalamo-cortical loop accompanied by increased oxidative stress (Cauli et al., 2009a, 2009b, 2006). Furthermore, clinical symptoms of HE typically include impaired motor performance or, in higher disease stages, mini-asterixis linked to altered electrophysiology in the thalamus (Butz et al., 2013, 2010; Timmermann et al., 2008, 2003). Other electrophysiological alterations include decreased cerebellar inhibition suggesting a gradual increase of GABAergic neurotransmission (Hassan et al., 2019) and slowed beta oscillations in the left motor cortex, which closely correlate with MRS-derived GABA+/Cr levels (Baumgarten et al., 2016). Finally, chemical exchange saturation transfer imaging in HE patients, which is sensitive to ammonia concentrations and microstructural cell alterations (Zöllner et al., 2019), as well as ^13^NH_3_-PET imaging (Keiding et al., 2006; Lockwood et al., 1991), suggested altered ammonia uptake in the cerebellum and thalamus. In summary, evidence from multiple modalities and disease models suggest that ammonia-induced changes of neurotransmission systems in the cerebello-thalamo-cortical system of HE patients are likely contributing to the occurrence of motor symptoms.

The goal of this study was therefore to investigate neurometabolic alterations in the cerebello-thalamo-cortical system of patients with HE. To this end, GABA-edited MRS was used to determine GABA+ levels in the cerebellum, thalamus, and motor cortex with the MEGA-PRESS sequence at 3T. In addition, MRS data were used to investigate disturbances in other neurometabolites. Finally, we assessed metabolite estimate differences between controls and patients at group level and determined their correlations with clinical metrics.

## Methods

### Study participants

35 participants were enrolled in the study, including 16 healthy age-matched controls, 3 minimal HE (mHE) patients, and 16 patients with clinically manifest HE (HE). Exclusion criteria for both controls and patients included psychiatric and neurological disease other than HE, severe intestinal disease, the use of any medication acting on the central nervous system, diagnosed peripheral/retinal neuropathy, and contra-indication for MR imaging such as pacemakers. If alcohol abuse was part of the medical history, the patient had to abstain from alcohol >= 4 weeks prior to enrollment. The study was performed in accordance with the current version of the Declaration of Helsinki (JAMA, 2013) and was approved by the local institutional review board (study number 5179R). All participants gave written informed consent before the examination.

### Grading and clinical metrics

HE severity grading according to *West-Haven* criteria (Ferenci et al., 2002; Kircheis et al., 2002) was performed by an expert physician (MSJ).

Clinical metrics included the critical flicker frequency (CFF) and fine motor performance with the timed pegboard test (tPEG). The CFF was included as it has been shown to be a reliable tool for HE diagnosis (Kircheis et al., 2002) and its quantitative assessment is able to reflect the continuous nature of HE symptom deterioration (Kircheis et al., 2014). CFF is typically around 40-44 Hz in healthy participants, and decreases steadily with HE severity, i.e. smaller CFF values indicate more pronounced disease severity. It was determined using portable goggles (NEVOlab, Maierhöfen, Germany). Fine motor performance was evaluated with a grooved pegboard test (Lafayette Instrument, Lafayette, IN, USA). Here, participants place pegs shaped like keys into a board with a 5 x 5 matrix of randomly oriented keyholes. The test is administered once for each hand and the scores are the sum of time to completion in seconds and errors for each hand defined as number of dropped pegs (Klove, 1963). The tPEG score is around 165 for healthy subjects between 55 and 70 (Heaton, 2004) and increases with increased deterioration of eye-hand coordination and motor speed as they occur in HE. All patients underwent blood testing to assess blood ammonia levels (NH_3_). Patients graded as ‘minimal HE’ underwent additional neuropsychological computer testing for diagnosis (Computer-based neuropsychometric tests from the Vienna Test System; Dr. Schuhfried GmbH, Mödling, Austria). The study cohort demographics are summarized in Table 1.

**Table 1.** Study cohort overview including all measured participants with graduation. Bold letters indicate p < 0.001 compared to the control group.

|  | mean (SD) |  |  |
| --- | --- | --- | --- |
|  | controls<br>n = 16 | mHE<br>n = 3 | HE<br>n = 16 |
| Age (years) | 65.8 (4.7) | 68.3 (3.8) | **66.6 (9.5)** |
| CFF (Hz) | 43.2 (2.4) | 38.5 (2.4) | **35.7 (2.2)** |
| tPEG score | 152.9 (152.5) | 195.3 (2.5) | **327.3 (285)** |
| Blood NH<sub>3</sub> (μg/dl) | - | 88.5 (40.3) | 97.3 (54.6) |

### Data acquisition

All MRS and MRI data were acquired on a clinical 3T scanner (Siemens MAGNETOM Prisma, Siemens Healthcare AG, Erlangen, Germany). The protocol included an anatomical T_1_-weighted scan for localization and segmentation purposes and three J-difference GABA-edited MEGA-PRESS (Mescher et al., 1998) acquisitions using *Siemens prototype sequence* (TE/TR = 68/1750 ms, NEX = 256, ‘ON’/’OFF’ editing pulse frequency = 1.9/7.5 ppm, editing pulse bandwidth = 82.4 Hz, spectral width = 2000 Hz, datapoints = 2048) with approximately 8 minutes of acquisition time per voxel. Water-unsuppressed scans (NEX = 16) with the same parameters were acquired for eddy current correction and water referencing. The MRS voxels were positioned in the right cerebellum (24 × 24 × 24 mm^3^ = 13.8 ml), the left thalamus/basal ganglia (30 (AP) × 35 (LR) × 25 (HF) mm^3^ = 26.25 ml), and the left motor cortex (30 × 30 × 30 mm^3^ = 27 ml) (see Figure 1). The cerebellar voxel was placed to include the dentate nucleus, the thalamic voxel was chosen to include the whole basal ganglia region to maximize the thalamic volume in the voxel (Dydak et al., 2011), and the cortical voxel was centered on the ‘hand knob’ and rotated to avoid the skull. The data were exported as .dat files for further processing.

**Figure 1.**
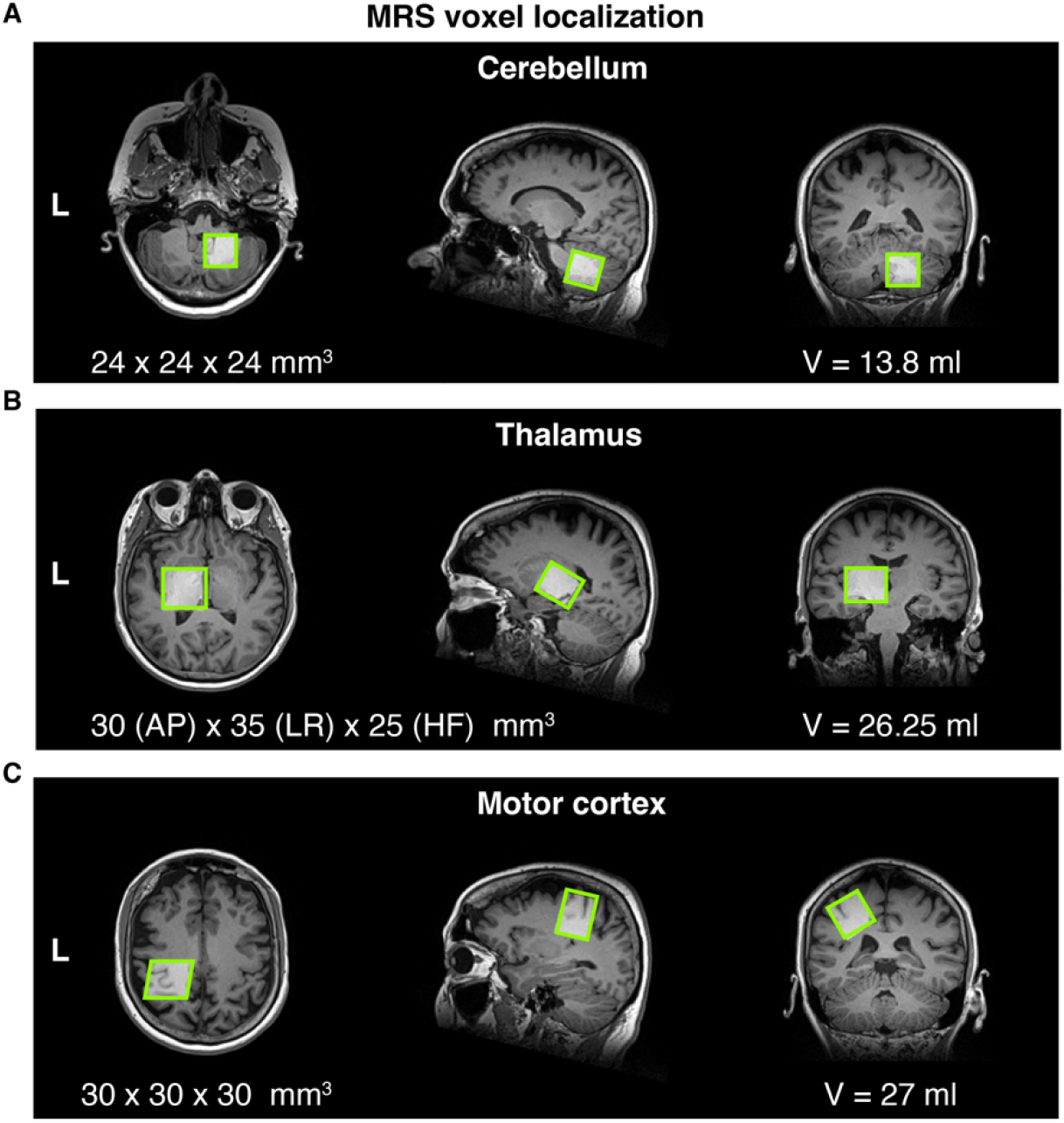
Localization of the GABA-edited MRS voxels from a representative subject. (A) Cerebellar voxel (B) Thalamus/basal ganglia voxel (C) Motor cortex voxel.

### Data Analysis

Two pipelines were used to analyze the GABA-edited difference and the un-edited (‘edit-OFF’) spectra. A summary of all experimental procedures following the ‘minimum reporting standards in MRS’ consensus paper (Lin et al., 2021) can be found in **Supplementary Material 1**.

### GABA-edited spectra

GABA was quantified from the J-difference-edited difference spectra using Gannet 3.1.2 (Edden et al., 2014), a MATLAB-based software specifically developed for this purpose. The processing included coil-combination, a probabilistic spectral registration approach for frequency-and-phase correction of the individual transients (Near et al., 2015), averaging of the transients, eddy-current correction using the water reference scan (Klose, 1990), zero-filling to 32768 data points, 3-Hz exponential line broadening, and removal of the residual water via Hankel singular value decomposition (HSVD) (Barkhuijsen et al., 1987). A three-Gaussian function with a non-linear baseline was used to model the 3-ppm GABA+ and 3.75-ppm Glx peaks of the resulting difference spectra, using non-linear least-squares fitting. The 3-ppm peak is referred to as GABA+, as the editing pulse has finite selectivity and is therefore co-editing macromolecular signals at 3 ppm with a coupling partner at 1.7 ppm, resulting in a composite GABA + macromolecule = GABA+ peak at 3-ppm with 40-60% of the signal originating from macromolecules (Choi et al., 2021; Petroff et al., 2001; Zöllner et al., 2021b).

### Edit-OFF spectra

The edit-OFF spectra were processed in Osprey v.2.3.0 (Oeltzschner et al., 2020), including probabilistic spectral registration for frequency-and-phase correction of the individual transients (Near et al., 2015), weighted averaging, eddy-current correction using the water reference scan (Klose, 1990), and HSVD water removal (Barkhuijsen et al., 1987). The data was then exported as .RAW files for quantification in LCModel v6.3 (Provencher, 2001). The basis set used for the linear-combination modeling of the edit-OFF spectra was generated using MRSCloud (Hui et al., 2022), a cloud-based toolbox to simulate basis sets using FID-A-derived matrix density simulations (Simpson et al., 2015). The basis set included 17 metabolites: aspartate, creatine, negative creatine methylene (-CrCH_2_ to account for water suppression/relaxation), GABA, glycerophosphorylcholine (GPC), glutathione (GSH), glutamine (Gln), glutamate (Glu), Myoinositol (mI), lactate, NAAG, NAA, scyllo-inositol, phosphoethanolamine (PE), and taurine. The macromolecules were included as 8 Gaussian basis functions (MM_0.91_, MM_1.19_, MM_1.38_, MM_1.65_, MM_2.03_, MM_2.27_, MM_2.6_, MM_3.01_, and MM_3.2_). The spectra were analyzed between 0.5 and 4 ppm, the baseline knot spacing parameter DKMNT was set to 0.4 ppm, and no further relaxation correction was employed in LCModel. For the motor cortex spectra, a GAP parameter between 0.95 and 1.95 was introduced to the LCModel fit to reduce the impact of lipid contamination artifacts (Provencher, 2020).

### Coregistration & Segmentation

The co-registration of the MRS volumes and the T_1_-weighted anatomical images, as well as the subsequent segmentation of the tissue composition, were carried out in the respective software (Gannet and Osprey). Both softwares use the ‘Coregister’ and ‘New Segment’ functions of SPM12 (Friston, 2007) to estimate gray matter (GM), white matter (WM), and cerebrospinal fluid (CSF) in the MRS volume. In addition, Osprey generates the forward deformation field to transform each voxel mask into MNI152 standard coordinate space, which was used to create across-subject voxel placement consistency maps for each MRS voxel.

### Quantification

All metabolite estimates were reported with respect to the total creatine (tCr = Cr + PCr) signal (metabolite-to-tCr ratio) and as water-referenced pseudo-absolute molal units (moles of metabolite per kg of solute water) corrected for partial volume effects according to the tissue composition of the individual MRS volumes and tissue-specific relaxation (Gasparovic et al., 2006). Both quantification approaches are commonly used in MRS, but each have potential sources of bias. Referencing to the internal creatine signal is sensitive to pathology-related creatine changes, including concentration and relaxation effects. In contrast, water-referenced methods are sensitive to changes in apparent water concentration and water relaxation. The effect is potentially amplified in subjects with HE as the literature suggests differences in several neuroimaging modalities, including evidence for relaxation and water content (Shah et al., 2008, 2003) potentially affecting the quantification. We therefore chose to report metabolite level estimates with both reference methods to minimize the risk of mis-interpreting changes to the reference compound.

### Quality metrics

Three quality metrics were applied to assess the spectral quality: visual inspection of the spectra, the full-width half-maximum (FWHM) and SNR estimates of the creatine peak in the edit-OFF spectra (Wilson et al., 2019), and the fit error of the 3-ppm GABA+ signal, defined as the fit amplitude divided by the standard deviation of the fit residual. Spectra with lipid contamination greater than twice the size of the NAA 2-ppm singlet were excluded.

### Statistics & Visualization

Group mean differences in the metabolite estimates were only assessed between healthy controls and the HE group, because the study cohort only included 3 mHE patients. The correlation analysis included the whole study cohort. Group mean differences were calculated using the lm() function of in R (Version 4.1.3). Similarly, a weighted partial correlation was used to assess associations between metabolite estimates and clinical metrics using the weights package (Pasek et al., 2021). Within-group correlations with clinical metrics were also evaluated for the healthy controls and the HE group. All *p*-values were adjusted using the Benjamini-Hochberg procedure (Benjamini and Hochberg, 1995) to account for false positives using an FDR rate of 5% as defined in the p.adjust function in R, including all comparisons for the GABA-edited (3 creatine-referenced group comparisons + 3 water-referenced group comparisons + 9 creatine correlations + 9 water-referenced correlations = 24) and edit-OFF (8 creatine referenced / 9 water-referenced group comparisons + 27 creatine referenced / 9 water-referenced correlations = 37) results. All statistics included age as a covariate and the metabolite-specific model uncertainty, defined as Cramér-Rao Lower Bound (CRLB), as a weighting factor to propagate the quantification uncertainty of the linear-combination model into the statistical model (Miller et al., 2017). The weighting factor was defined as the metabolite-specific inverse of the absolute CRLB (Fowler et al., 2021).

Metabolite estimates are visualized as raincloud plots (Allen et al., 2019) with mean, median, 25^th,^ and 75^th^ percentile range, individual estimates, and smoothed distributions. Additionally, bar plots with standard deviation and individual estimates generate a more compact visualization. Correlations between metabolite estimates and clinical metrics are visualized in a correlation matrix which includes the weighted partial correlation coefficients *r*. A color-coded (blue = positive, red = negative correlation coefficient) ellipsoid indicates significant associations after correction for multiple comparisons, with higher associations represented by a narrower ellipsoid, i.e., perfect associations (r=1) would result in a line and no association (r = 0) in a circle. Additionally, the weighted partial correlation coefficients *r* of the significant associates are marked in bold while non-significant are reported in gray. All plots and statistics are generated in R (Version 4.1.3) in RStudio (Version 2022.02.01, Build 461, RStudio Inc.) using SpecVis (Zöllner et al., 2021a), an open-source package to visualize linear-combination modeling results with the ggplot2 package (Wickham, 2009). All scripts are publicly available on the SpecVis GitHub repository (https://github.com/HJZollner/SpecVis).

## Results

The remaining datasets after quality control included 30 spectra from the cerebellum, 34 spectra from the thalamus, and 28 spectra from the motor cortex. 5 datasets from the cerebellum were removed due to out-of-volume (OOV) echoes (1 control and 3 HE patients) or unreasonably high GABA+ estimates (1 HE patient with 2.4-fold increased GABA+ estimates). One dataset from the thalamus were removed due to OOV echoes (1 HE patient). For the motor cortex HE patients cancelled during the acquisition of this voxel and 5 datasets were removed due to a lipid contamination (2 controls, 1 mHE, and 1 HE patient). A summary of the quality metrics of the remaining spectra can be found in Table 2. GABA fit errors for the GABA-edited spectra and metabolite CRLBs for the edit-OFF spectra can be found in **Supplementary Material 2**.

**Table 2.** Summary of the spectral quality metrics and voxel tissue composition across the study cohort. No systematic difference in spectral quality was found between groups. The thalamus voxel contained less WM and more CSF in HE patients than in controls. Bold = significantly different (p < 0.05) to controls.

|  | **mean (SD)** |  |  |
| --- | --- | --- | --- |
|  | controls | mHE | HE |
| **Cerebellum** |  |  |  |
| n | 15 | 3 | 12 |
| SNR | 124.8 (31.2) | 139.9 (9.9) | 138.3 (23.6) |
| FWHM (Hz) | 8.6 (0.9) | 8.3 (0.1) | 8.3 (0.6) |
| **Thalamus** |  |  |  |
| n | 16 | 3 | 15 |
| SNR | 128.7 (28.1) | 144.9 (32.3) | 109.6 (22.9) |
| FWHM (Hz) | 11.7 (1.8) | 11 (1.7) | 12.7 (1.4) |
| **Motor cortex** |  |  |  |
| n | 14 | 2 | 12 |
| SNR | 258 (62.3) | 236.7 (27.3) | 220.4 (49.6) |
| FWHM (Hz) | 7.7 (0.3) | 7.47 (0.3) | 8.2 (1.0) |
| **Cerebellum** |  |  |  |
| GM (%) | 49 (5) | 47 (5) | 49 (6) |
| WM (%) | 49 (5) | 50 (5) | 47 (9) |
| CSF (%) | 2 (1) | 4 (3) | 4 (4) |
| **Thalamus** |  |  |  |
| GM (%) | 31 (3) | 34 (6) | 31 (5) |
| *WM (%)* | 60 (4) | 57 (1) | **55 (5)** |
| *CSF (%)* | 9 (3) | 9 (6) | **14 (5)** |
| **Motor cortex** |  |  |  |
| *GM (%)* | 25 (3) | 23 (0) | 25 (3) |
| *WM (%)* | 63 (4) | 64 (0) | 63 (4) |
| *CSF (%)* | 13 (2) | 13 (1) | 13 (4) |

The thalamus voxel contained less WM and more CSF in HE patients than in controls, indicating increased ventricular atrophy. All other tissue characteristics did not differ significantly between groups.

The spectra of all remaining participants are presented in Figure 2 **A**. Mean spectra and standard deviation of the control and HE group are also shown. The overall data quality is good with acceptable linewidth and SNR (Wilson et al., 2019), and there were no systematic differences in data quality and appearance between both groups. Compared to the cerebellum and thalamus, lipid contamination can be observed in the 0.5 to 1.95 ppm region of the edit-OFF motor cortex spectra. The frequency region between 0.95 and 1.95 was excluded during the modeling process to reduce the impact of the artifacts on the overall quantification. In addition, a substantial signal increase in the 3.8 ppm region of the GABA-edited difference spectra can be observed for the HE group indicating elevated levels of Gln, GSH, and Glu (see arrows). The edit-OFF spectra reveal the HE-typical metabolic patterns revealing increased signals in the region for Gln, GSH, Glu, and reduced signal for mI and tCho (see arrows in Fig 2).

**Figure 2.**
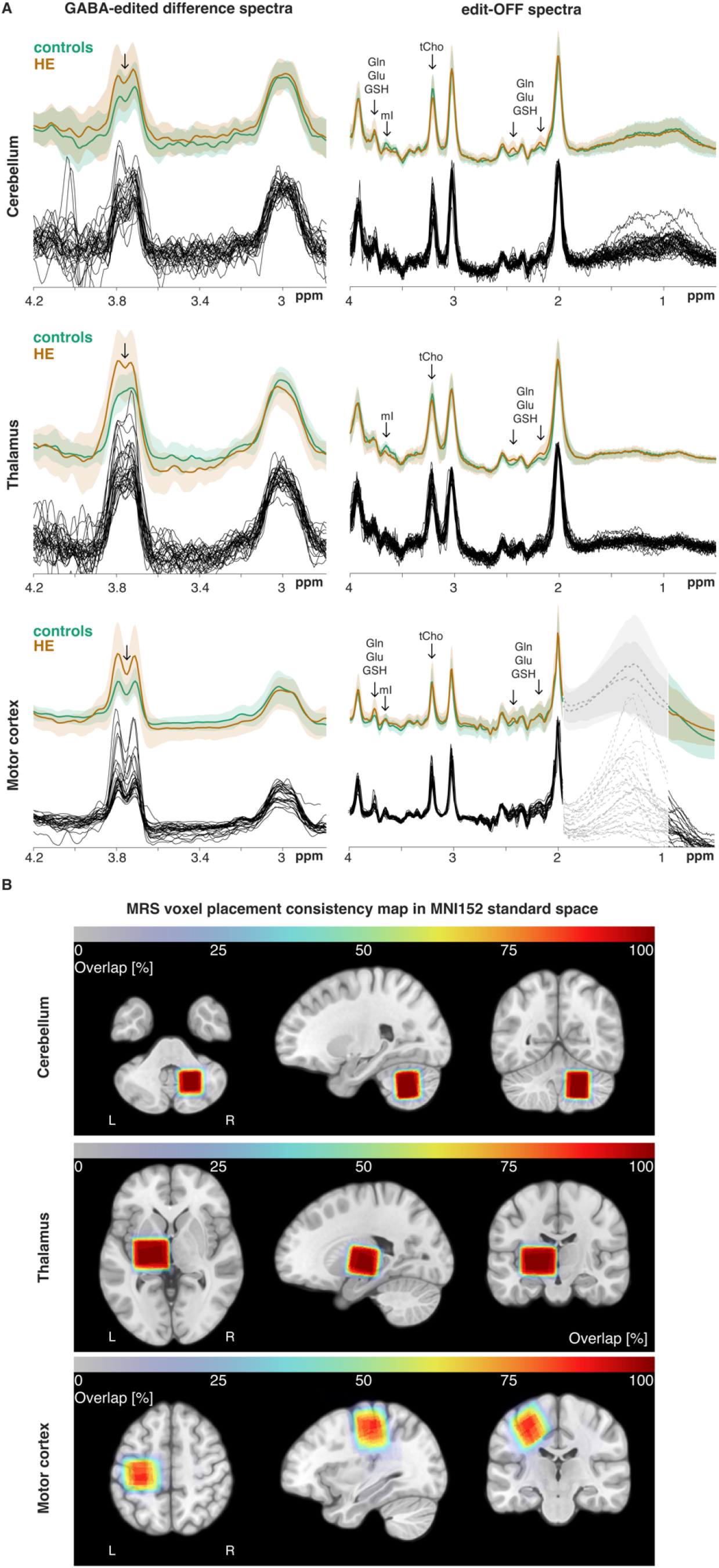
Overview of the MRS data. (A) Individual GABA-edited and edit-OFF spectra from the cerebellum, thalamus, and motor cortex. Additionally, the control and HE groups’ mean and standard deviation spectra are shown for all spectra. Data quality did not differ between groups. The substantial increase in the Glx signal of the GABA-edited spectra is visible in the HE group, clearly reflecting the strong characteristic Gln increase in HE. The frequency region excluded during the modeling is indicated in gray. (B) Voxel placement consistency maps for all three MRS voxels overlayed on the MNI152 template show high reproducibility for the cerebellum and thalamus voxel. In the course of the study, the motor cortex voxel was increasingly moved further away from the skull to reduce lipid contamination, resulting in higher placement variability.

The cerebellum and thalamus voxels were placed with excellent consistency between subjects (Figure 2B). Motor cortex voxel placement was more variable as more conservative voxel placement was established over the course of the study to reduce lipid contamination. This caused slightly smaller, but still very good overlap compared to the other MRS voxels.

### GABA+ results

GABA+ estimates for all three voxels are summarized in Figure 3. The Cr-referenced GABA+ (Figure 3 A-C) was significantly increased in the cerebellum and significantly decreased in the motor cortex in HE patients. No changes were found in the thalamus. Additionally, GABA+ levels in the motor cortex were significantly correlated with CFF (positive) and total tPEG score (negative) (Figure 3C). Similarly, the water-referenced GABA+ levels were elevated in the cerebellum and reduced in the motor cortex of patients with HE (Figure 3D), confirming the Cr-referenced estimates. Compared to the creatine-referenced GABA+ levels, an even stronger association with the clinical metrics was found (Figure 3F). The quantitative results of the full study cohort are summarized in Table 3. No significant association with blood ammonia levels was found for any reference method. However, this could also be related to fewer participants with available blood test results. Further, no within-group correlations were found between both GABA+ and any of the reported clinical metrics.

**Figure 3.**
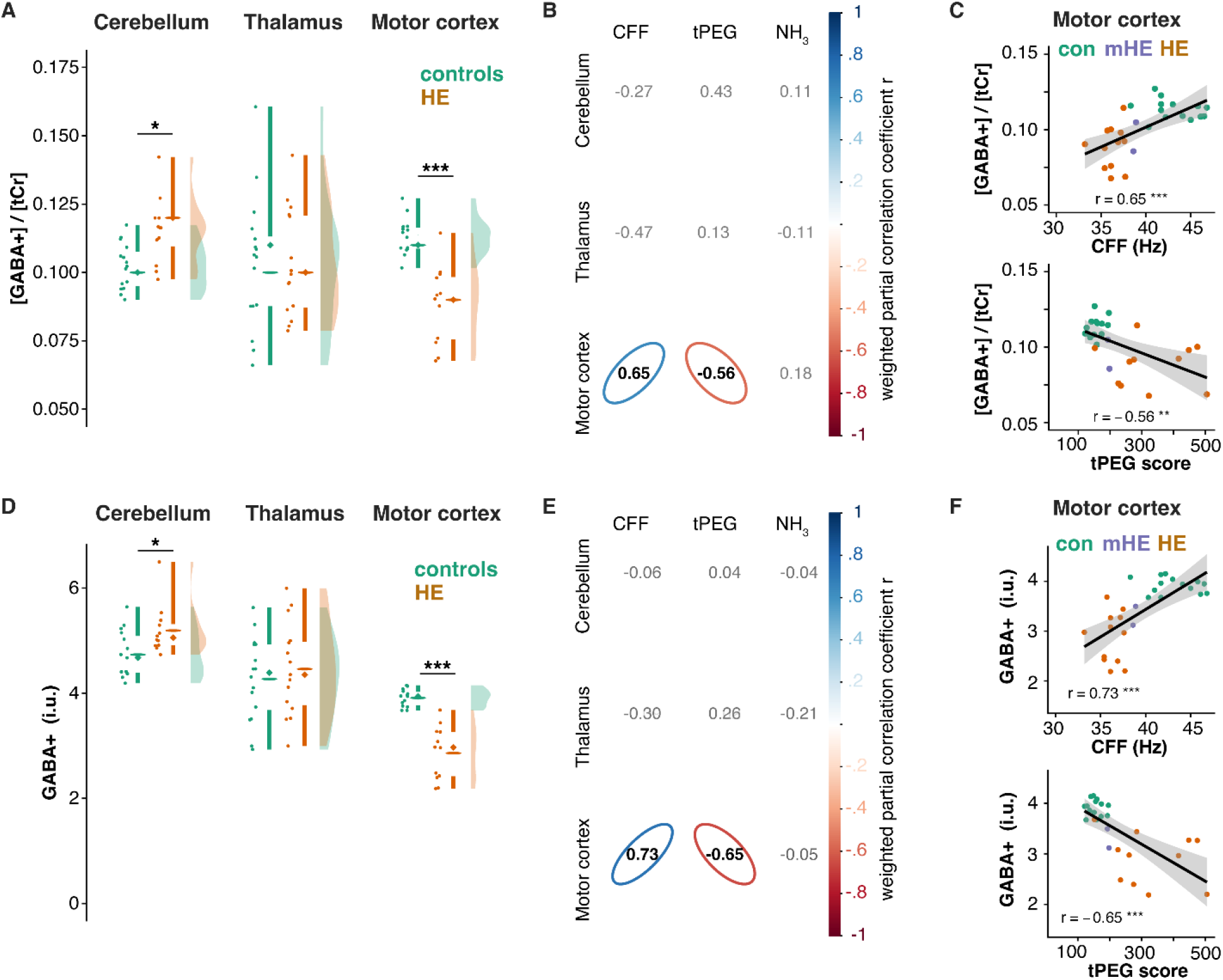
Summary GABA+ estimates from the GABA-edited spectra. (A) – (C) Creatine-referenced GABA+ estimates, correlation with clinical metrics for all voxels, and significant associations from the motor cortex MRS voxel. GABA+/tCr estimates were significantly increased in the cerebellum of HE patients and significantly reduced in the motor cortex. GABA+ estimates correlated with CFF and tPEG score values. (D) – (F) Water-referenced GABA+ estimates and correlations with clinical metrics reproduce the creatine-referenced findings with a stronger association with the clinical metrics. The raincloud plots include mean (horizontal line), median (diamond), 25^th^ and 75^th^ percentile (vertical lines), individual estimates, and smoothed distributions. Comparisons and correlations were corrected for multiple comparisons using the Benjamini-Hochberg procedure with an FDR of 0.05.

**Table 3.**
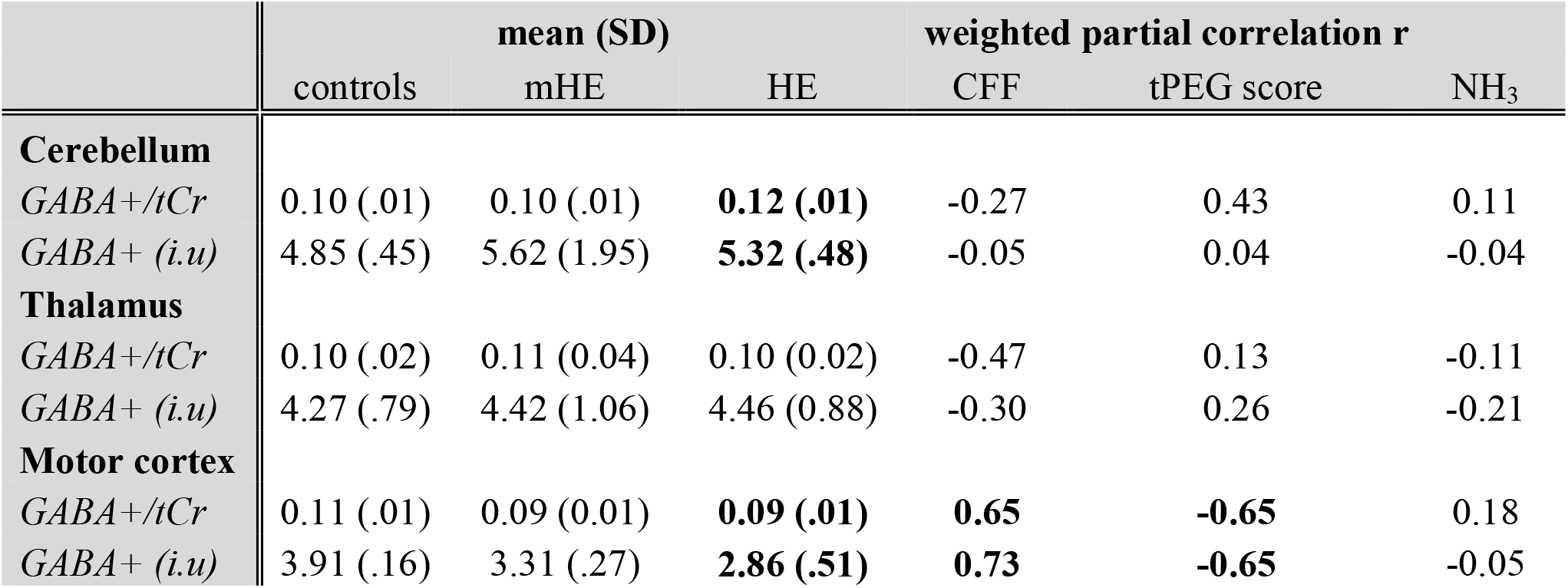
Summary GABA+ estimates from the GABA-edited spectra. Significant differences between the healthy control and the HE group and significant correlations between the clinical metrics and the GABA+ levels are indicated in bold. Comparisons and correlations were corrected for multiple comparisons using the Benjamini-Hochberg procedure with an FDR of 0.05.

### Edit-OFF spectra results

The analysis of the edit-OFF spectra is summarized in Figure 4, including metabolites typically affected in HE. A summary of the statistics for all quantified metabolites can be found in **Supplementary Materials 3, 4**, and **5**. Characteristic HE-typical metabolite patterns are observed in all MRS voxels, i.e. significantly increased Gln and reduced mI levels for both referencing methods (Figure 4 A & C). Additionally, glutamate levels were significantly increased in the thalamus. However, this finding may rather arise as a modeling artefact resulting from poorer spectral resolution (higher FWHM) in the thalamus and, therefore, more challenging separation of the glutamine and glutamate signals with LCM. Generally, lower relative CRLBs were found for the glutamine estimates in HE patients (12.4 ± 6.7 %) due to higher concentrations, which results in more reliable estimation than in healthy controls (24.3 ± 5.6 %). GSH levels in HE were increased in the thalamus and motor cortex for the Cr-referenced estimates (Figure 4A) and the water-referenced thalamus estimates (Figure 4C). Total choline was reduced in HE in the creatine-referenced levels in the thalamus (Figure 4A) and in all three MRS voxels for water-referenced estimates (Figure 4C).

**Figure 4.**
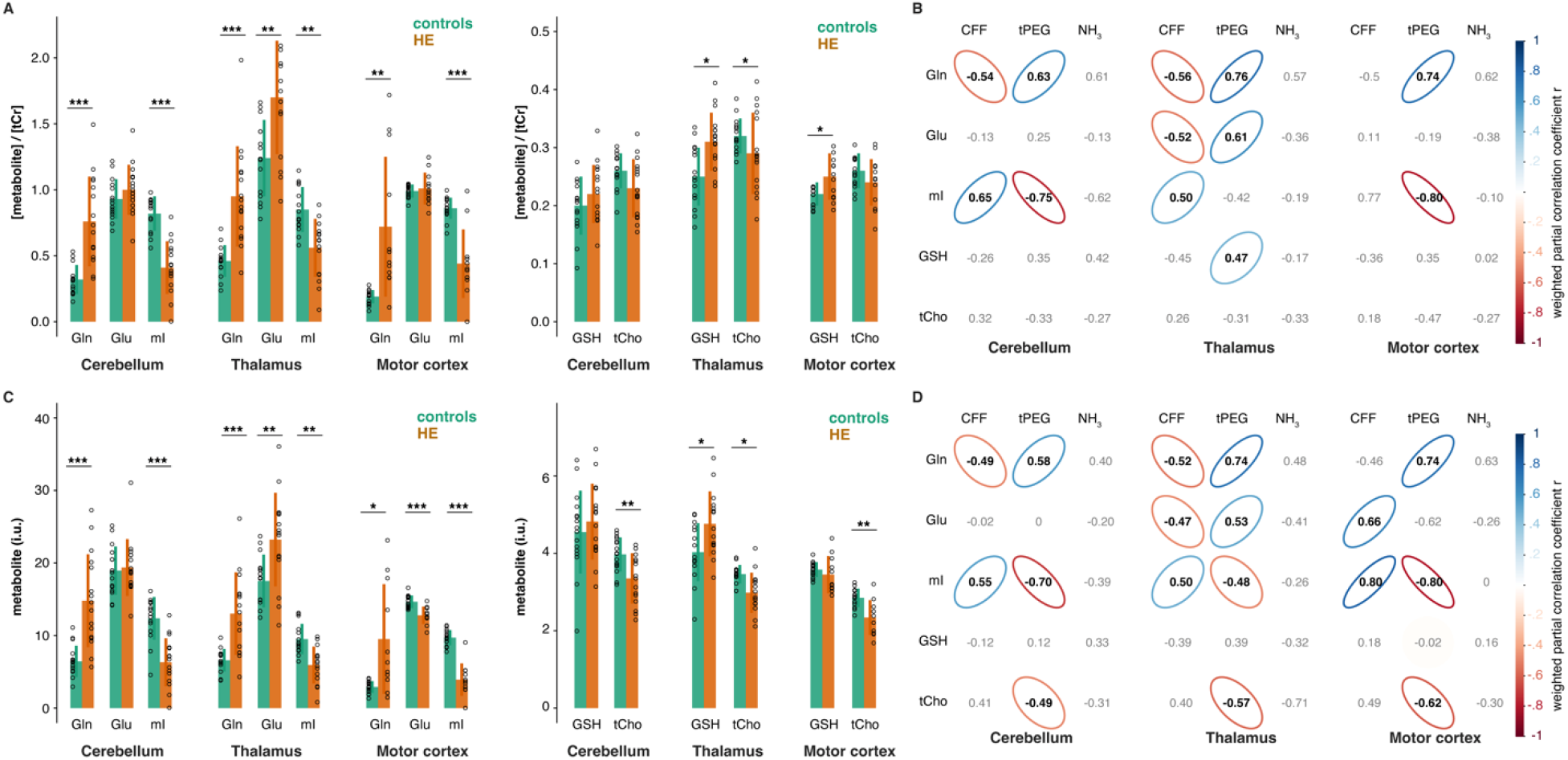
Summary of HE metabolite profiles from the edit-OFF spectra. (A) Cr-referenced metabolite estimates (mean + SD & individual estimates) for all MRS voxels, including glutamine, glutamate, Myo-inositol, glutathione, and total choline. (B) Correlations between Cr-referenced metabolite estimates and clinical metrics. (C) Water-referenced metabolite estimates for all voxels. (D) Correlations between water-referenced metabolite estiamtes and clinical metrics. The HE-typical metabolite profile (increased Gln accompanied by mI reduction) was consistently found in all MRS voxels. Additionally, increased GSH (thalamus & motor cortex) and reduced tCho (cerebellum, thalamus & motor cortex) were found in some voxels. Metabolite estimates correlated strongly with clinical metrics. Glutamine vs. blood ammonia levels correlation coefficients were high in all MRS voxels, but not significant after multiple comparison corrections.

Strong correlations with clinical metrics were found for Gln in the cerebellum (CFF & tPEG for both reference methods), thalamus (CFF & tPEG for both reference methods), and motor cortex (tPEG for both reference methods). Thalamic Glu correlated strongly with CFF and tPEG scores for both reference methods and water-referenced levels in the motor cortex. Cr-referenced mI correlated with CFF in the cerebellum and thalamus and with tPEG scores in the cerebellum and motor cortex. Water-referenced mI correlated with CFF and tPEG scores in all three MRS voxels. Further, Cr-referenced GSH levels correlated with tPEG scores in the thalamus MRS voxel, and water-referenced total choline levels correlated with tPEG scores in all three MRS voxels. Correlations with blood ammonia levels were not significant after multiple comparison corrections, although the correlation coefficients for Gln was high (Figure 4B & D). This is certainly caused by the limited range of blood ammonia measures, as blood tests were only conducted in the patient groups. Generally, group comparisons and correlations were highly consistent between MRS reference methods. These metabolic patterns also agree strongly with many earlier findings in MRS HE literature (Häussinger et al., 1994; Kreis et al., 1991; Miese et al., 2006; Zeng et al., 2020).

Further, water-referenced tNAA was significantly reduced in HE in the thalamus and motor cortex, and tCr was reduced in HE for all three MRS voxels (**Supplementary Material 3** and **4**). This tCr group difference is remarkable and additionally supported the use of two different reference methods. Water-referenced tNAA levels correlated with blood ammonia levels in the thalamus, and tCr levels correlated with tPEG scores in the cerebellum and CFF in the motor cortex (**Supplementary Material 5**).

### Explorative results

As described in the methods, additional metabolites typically found in the healthy human brain are routinely included in MRS basis sets. However, the reliable quantification of those metabolites with LCM is challenging due to low concentrations and spectral overlap. Significant pathological changes that we found for some metabolites, aspartate (Asp) and scyllo-inositol (sI), must therefore be interpreted with great care. We included metabolite-specific model uncertainty (given by CRLB) in the final statistics to account for this. Figure 5 summarizes the aspartate estimates. Cr-referenced Asp estimates were significantly increased in HE in all three MRS voxels (Figure 5A) and in the cerebellum and thalamus for water-referenced estimates (Figure 5B). Cr-referenced estimates correlated with CFF for all three voxels and in the cerebellum and thalamus for the water-referenced estimates. Additionally, thalamic Asp correlated with the total PEG score.

**Figure 5.**
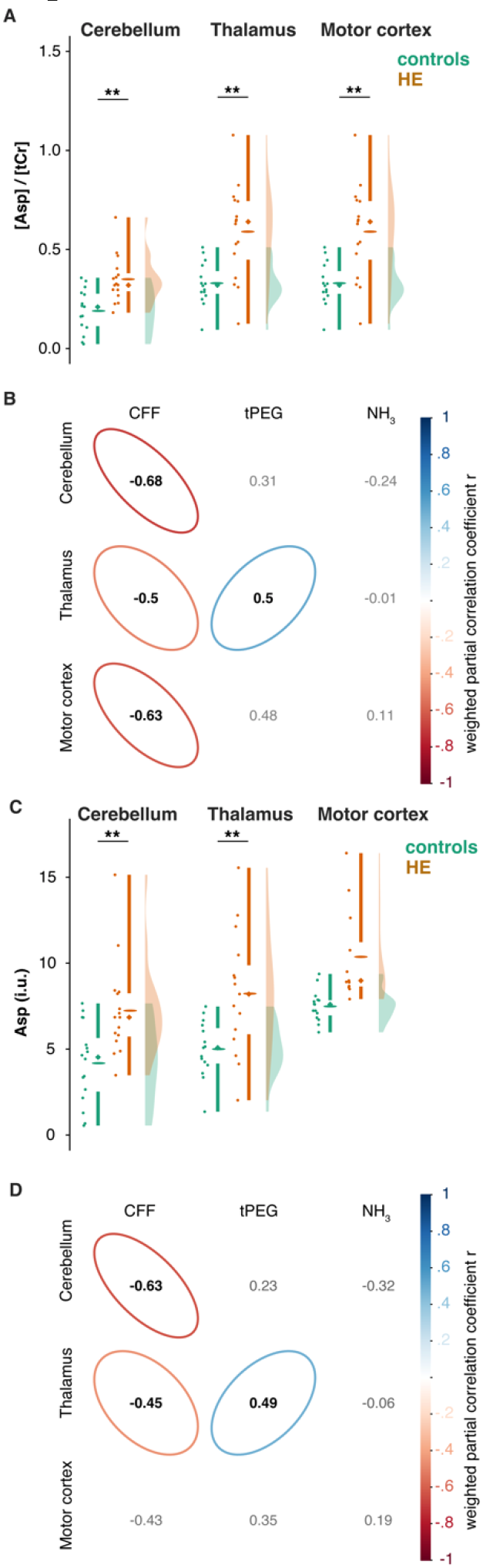
Explorative analysis of aspartate estimates in the edit-OFF spectra. (A) Creatine-referenced aspartate estimates for all MRS voxels. (B) Correlations between creatine-referenced aspartate estimates and clinical metrics. (C) Water-referenced aspartate estimates for all voxels. (D) Correlations between water-referenced aspartate estimates and clinical metrics. Aspartate estimates were significantly increased in all MRS voxels in the HE group and correlated strongly with the CFF. Again, a strong correlation with the blood ammonia levels was found that was not significant after multiple comparison corrections.

Further, sI estimates were significantly reduced in the motor cortex, where they additionally correlated with CFF and total PEG score (**Supplementary Material 3**, **4**, and **5**).

## Discussion

We have investigated the cerebello-thalamo-cortical metabolism of patients with HE using GABA-edited MRS at 3T. The aims of the study were: 1) to investigate GABA levels in the cerebello-thalamo-cortical system, 2) to translate and scrutinize findings from HE animal models to humans (Cauli et al., 2009a, 2009b), 3) to substantiate electrophysiological observations of GABAergic tone with complementary MRS measurement (Hassan et al., 2019), and 4) to test whether HE-typical metabolic patterns are consistent across the cerebello-thalamo-cortical system (Zeng et al., 2020).

The key findings of our study are:

- GABA+ levels in patients with HE were significantly increased in the cerebellum and decreased in the motor cortex. Cortical GABA+ levels correlated strongly with CFF and tPEG scores. Thalamic GABA+ levels did not differ between controls and HE patients.
- The characteristic metabolic patterns of HE (increased Gln and GSH; decreased mI and tCho) appeared in all three regions, and a strong association of this pattern with clinical metrics was found.
- Total creatine was reduced in all three regions; tNAA was reduced in the thalamus and motor cortex.
- Aspartate levels were significantly higher in HE patients than controls in all three regions and were strongly associated with the clinical metrics. Further, scyllo-inositol levels were reduced in the motor cortex and associated with disease severity.

To the authors’ best knowledge, this is the first study to investigate in-vivo GABA levels in all three key regions of the human cerebello-thalamo-cortical system using GABA-edited MRS. We found substantially altered in-vivo GABA levels, which were additionally linked to clinical metrics and accompanied by a general metabolic disturbance in the motor system. These results support earlier reports of increased extracellular GABA levels in the cerebellum and ventromedial thalamus in a hyperammonemia rat model accompanied by reduced cortical GABA levels (Cauli et al., 2009a). We found increased GABA+ levels in the cerebellum and decreased levels in the motor cortex, although thalamic levels did not differ significantly between groups.

There are several interpretations for this mismatch: (i) GABA-edited MRS measures the combined intra- and extracellular GABA concentration, deriving a ‘bulk’ tissue concentration across the whole MRS voxel. Subtle changes in extracellular GABA levels (Rae, 2014; Stagg et al., 2011) may be obscured by the much greater intracellular GABA pool. (ii) The MRS voxel used in this study included other basal ganglia structures that may also be (differentially) affected by HE. This includes pronounced changes in T_1_ relaxation (Shah et al., 2003), chemical exchange saturation transfer (Zöllner et al., 2019), magnetization transfer (Miese et al., 2006, 2009), and possibly water content (Oeltzschner et al., 2016; Shah et al., 2008). These changes may confound spectral quality and quantification accuracy leading to the overall higher variance in the GABA+ estimates that we observed in the thalamus, and therefore diminished statistical power for a constant sample size. (iii) The changes reported in the animal model were limited to the ventromedial thalamus and not observed in the mediodorsal thalamus (Cauli et al., 2009a), both of which are included in the MRS voxel. Additionally, the observed changes in the animal model were less pronounced in the ventromedial thalamus (1.6-fold increase) compared to the cerebellum (1.9-fold increase).

### GABA MRS in HE

Literature on in-vivo GABA levels measured with MRS to investigate HE is relatively scarce. Early applications reported reduced GABA + homocarnosine levels in the occipital region using GABA-edited MRS (Behar et al., 1999), which was later replicated (Oeltzschner et al., 2015). Another study found increased GABA+ levels in the left striatum of liver cirrhosis patients and a correlation with psychometric hepatic encephalopathy scores (Zupan et al., 2019). In contrast to our earlier study, (Oeltzschner et al., 2015), we found significantly reduced GABA+ in the motor cortex with strong associations with clinical metrics. In this study, we used longer repetition times (1750 *vs*. 1500 ms), a higher number of excitations (256 *vs.* 192 NEX), a higher number of datapoints (2048 *vs.* 1024), new state-of-the-art processing (probabilistic spectral registration *vs.* spectral registration), and an improved model to quantify the 3-ppm GABA+ peak (3 Gaussian & non-linear baseline vs. 1 Gaussian &linear baseline). Enhanced data quality and robustness of quantification may have improved our ability to resolve sensorimotor GABA+ changes. Studies using 2D correlation spectroscopy did not report significant differences in GABA levels in mHE patients in the frontal and occipital lobe (Singhal et al., 2010) and the anterior cingulate cortex (Binesh et al., 2005). However, these studies were conducted at 1.5 T and suffered from a highly variable estimation, diminishing the statistical power to find subtle concentration changes, which are generally likely to be more pronounced in than in minimal HE.

### MRS in HE

Many studies have used conventional and advanced MRS to investigate HE, summarized in a recent meta-analysis (Zeng et al., 2020). MRS has been proposed as a surrogate diagnostic marker that may outperform other methods in identifying mHE and monitoring the effectiveness of therapeutic strategies (Hermann et al., 2019). The meta-analysis of 31 studies found consistent metabolic patterns that we confirmed in the present study, i.e. increased Glx (glutamine + glutamate) and reduced myo-inositol levels in parietal regions and basal ganglia of HE patients, and reduced choline levels in the basal ganglia of HE patients. In addition, we found increased glutamate levels in the thalamus, which are difficult to interpret due to limited literature with separate glutamine and glutamate results. They may also arise as a result of the poorer separability of both compounds in the thalamus due to higher linewidth in this technically challenging region. The pathogenesis of HE has been associated with inflammation, neurotoxicity, and oxidative stress (Häussinger et al., 2021; Simicic et al., 2022). Glutathione is a cellular antioxidant marker in the body’s response to these stressors. Several cell and animal studies have reported increased total glutathione levels (reduced + oxidized glutathione) under hyperammonemic conditions (Murthy et al., 2000; Węgrzynowicz et al., 2007). While optimized protocols for the MRS detection of GSH or even simultaneous detection of GABA and GSH exist (Saleh et al., 2016; Terpstra et al., 2003), it may also be reliably detected in the edit-OFF spectrum of a GABA-edited experiment (Michels et al., 2014). Our exploratory analysis also found increased GSH levels in the motor cortex, confirming the findings of our earlier study (Oeltzschner et al., 2016), as well as elevated levels in the thalamus. Interpreting these results with regards to the partial volume effect of higher white matter fractions in these regions (motor cortex: 63%; thalamus: 57%) compared to the cerebellum (48%) could imply that the majority of the antioxidative GSH response takes place in white matter astrocytes. However, this interpretation contrasts studies on cortical astrocytes and cortical regions in rodents implying GSH synthetization to predominantly take place in gray matter. Therefore, the variability in the GSH levels is more likely to be region-specific alterations in the response mechanism similarly observed in region-specific changes of neurotransmitter systems in HE (Cauli et al., 2009b, 2009a, 2006). Studies employing edited or ultra-high field strength MRSI can improve the resolution of the small GSH signal and may help elucidate the region specificity of GSH levels and underlying oxidative stress mechanisms.

Findings regarding the total NAA level in HE remain inconclusive in the literature (Zeng et al., 2020). tNAA is generally considered a marker for neuronal integrity (Rae, 2014). However, HE is frequently described as a glial syndrome; therefore, an absence of changes is usually explained by this definition. In our study, we found reduced tNAA in the motor cortex and thalamus possibly related to HE-induced brain atrophy, especially as we found reduced white matter fractions in the thalamus of HE patients. While no such changes were found in the motor cortex, reduced levels may also be an earlier marker of neuronal loss, which has not precipitated as MR-visible atrophy, but is already observable at the biochemical level. Another interesting finding in our study is the systematic reduction of total creatine. Literature integration of these findings is challenging as many MRS studies use total creatine as a reference. However, they support animal studies reporting reduced thalamic creatine levels (measured with water-referenced MRS) (Cao et al., 2018). Total creatine is often interpreted as a marker for energy metabolism. Animal studies suggest decreased creatine kinase activity in the cerebellum and cortex affected by the oxidative stress response in HE (Pacheco et al., 2009). This indicates that water-referenced total creatine levels may add an additional marker for HE severity, although the specificity of this mechanism in HE needs to be further investigated.

The increase in aspartate levels found in our study and their close link to clinical metrics are novel findings. However, these preliminary findings must be interpreted with care since Asp is a low-concentration compound that typically requires advanced techniques for its reliable detection at a clinical field strength (Chan et al., 2017; Menshchikov et al., 2020, 2019). Studies at 1.5 T employing 2D correlation spectroscopy for more robust separation of j-coupled metabolite found a trend-level Asp increase in HE and a correlation with clinical metrics (Binesh et al., 2005; Singhal et al., 2010). These findings could be linked to disrupted glutamatergic neurotransmission, also visible in the altered glutamate levels found in this study, and the linked transamination of glutamate and aspartate. Further, aspartate plays a crucial role in the malate-aspartate shuttle (Rao and Norenberg, 2001) transporting glutamate into the mitochondria in exchange for matrix aspartate (Palmieri et al., 2001). Therefore, it may be a marker for disturbances in the energy metabolism and mitochondrial dysfunction (Rao and Norenberg, 2001). Lastly, we found decreased scyllo-inositol (sI) levels, which is a stereoisomer of myo-inositol. Early MRS studies suggested unquantified changes in a resonance at 3.35 ppm by creating a difference spectrum between healthy controls and HE patients (Kreis et al., 1991), which has later been assigned to sI and was repeatedly found in ex vivo tissue MRS of the cortical gray matter of HE patients at 7.8 T (Lien et al., 1994). These changes are suggested to occur in parallel with myoinositol reduction and seem limited to the gray matter as implied by our results. Another study in patients with chronic alcoholism found significant increased sI levels that were preceding the development of symptomatic hepatic encephalopathy (Viola et al., 2004).

### Other neurological findings in the cerebello-thalamo-cortical system

The general involvement of the cerebello-thalamo-cortical system in the neurophysiology and clinical symptoms has been demonstrated in several studies, including slowed oscillatory coupling related to mini-asterixis (Timmermann et al., 2008, 2003), altered functional connectivity in mHE (Qi et al., 2013), and neuroinflammatory response resulting in T_1_ relaxation changes (Shah et al., 2003). Sensorimotor deficits are a common symptom of HE and further support involvement of the motor system (Butz et al., 2010). Especially cerebellar damage is linked to such deficits and slower upper limb movements (Butz et al., 2010; Manto et al., 2012; Miall et al., 2007) substantiating the close association of the tPEG score and altered metabolism in the motor system presented here. In a previous study, we found altered amide-proton-transfer-weighted chemical exchange saturation transfer in the cerebellum of HE patients, which is likely sensitive to brain ammonia levels (Zöllner et al., 2019, 2018). The present study provides further evidence of altered cerebellar metabolism. Most notably, we have previously demonstrated decreased cerebellar inhibition using transcranial magnetic stimulation (TMS), suggesting a gradual increase of GABAergic neurotransmission in HE (Hassan et al., 2019). The increased cerebellar GABA+ levels in HE patients that we report in this study serve as a complementary piece of evidence to further support the link between cerebellar HE metabolism and functional symptom emergence.

### MRS methodological aspects

MRS literature suggests improved statistical power of in vivo MRS studies when a theoretically sound propagation of variable uncertainty is used (Miller et al., 2017). The actual application of those methods is, however, very sparse (Fowler et al., 2021). Here, we implemented a framework to include quantification uncertainty from Cramer-Rao lower bounds into the statistical analysis. The CRLBs were added as a weighting factor of the subject-specific data points for the group and correlation analysis to account for quantification uncertainty and increase the robustness of the statistics. The implementations are freely available in SpecVis (https://github.com/HJZollner/SpecVis).

Reporting both the creatine- and water-referenced estimates increased the transparency of this study and reduces possible referencing bias, an approach justified when we actually observed group differences in total creatine levels. Reporting water-referenced estimates with full tissue-volume and relaxation correction is now the expert consensus recommended method for virtually all MRS methods (Near et al., 2020). However, this approach – just like creatine referencing – relies on assumptions regarding water visibility and relaxation contents, and these assumptions may become invalid under certain pathological conditions, e.g. altered brain water content in HE. Differences in tissue composition could be accounted for by normalizing the results to the average composition of the healthy control group and possibly further including assumptions about gray and white matter metabolite concentrations (Harris et al., 2015). However, this further dilutes the interpretability of the pseudo-molal concentration units. Another approach is to calculate a combined reference metric of multiple metabolites (Larsen et al., 2021) or the use of external reference signals which are technically challenging to implement (Akoka and Trierweiler, 2002; Barantin et al., 1997).

### Limitations

It must be noted that the correlation analysis is limited by the low number of mHE patients (n = 3) and is therefore mainly driven by group differences. The data lacks a substantial portion of estimates from mHE patients to further substantiate the linear associations. This was further exacerbated by the fact that blood tests were only performed on HE patients. Within-group correlations were only significant for tNAA levels in the thalamus and blood ammonia levels due to the high number of for multiple comparison corrections.

In this study, we used a standard GABA-edited MRS scheme which results in substantial coediting of MM (Choi et al., 2021), leading to a composite peak of GABA and MM at 3 ppm. Both components are not separable with conventional acquisition or modeling methods. Therefore, we assumed the MM contribution to be stable between healthy controls and patients, and assume any changes of the composite signal to be driven by GABA. While there are methods to reduce the contamination of co-edited MM (Henry et al., 2001), those methods are extremely susceptible to subject movement and frequency drift.

While abundant care was taken during the positioning of the motor cortex MRS voxel, substantial lipid artifacts were present in several spectra, potentially due to movement of the participant, as the motor cortex voxel was acquired at the end of the protocol. There is no literature on the impact of fat contamination on editing efficiency and how to best account for them during processing and quantification. We used strict outlier removal criteria and probabilistic spectral registration to reduce the impact on the processing. The optimization of the Gaussian model for the GABA-edited difference spectra is not directly affected as the peak is outside of the model region. For the edit-OFF, we used the ‘GAP’ parameter in LCModel to reduce the impact during the optimization.

The separability of glutamine and glutamate, as well as aspartate and scyllo-inositol for LCM of 68-ms edited-OFF spectra at 3 T may be limited. We found reasonable CRLBs for the glutamine estimates (healthy controls (24.3 ± 5.6 %) and HE patients (12.4 ± 6.7 %), more details in **Supplementary Material 2**) compared to glutamate estimates in optimized STEAM acquisitions with high separability (Wijtenburg and Knight-Scott, 2011), suggesting that quantification may be feasible, but this has not been independently validated. Additionally, we accounted for the model uncertainty by propagating it into the statistical analysis. The higher variance in the glutamine estimates in healthy controls is mainly due to lower estimates, compared to the substantial increase in HE patients. Better separability of those metabolites may be achieved by employing other editing techniques (Oeltzschner et al., 2019), 2D correlation spectroscopy (Binesh et al., 2005; Singhal et al., 2010), or higher field strength to increase confidence in the reported findings.

Finally, changes in the tissue relaxation properties are associated with HE, with possible changes in water content or changes in the brain’s microstructure contributing to these changes. Therefore, metabolite and water relaxation times may also be affected by HE. However, current MRS techniques do not allow for the separation of concentration and relaxation time changes, possibly leading to systematic bias.

## Conclusion

We found alterations in the GABAergic neurotransmitter system in the cerebellum and motor cortex in HE. These changes were accompanied by a general disturbance of osmolytes and oxidative stress markers in the cerebello-thalamo-cortical system. These disturbances are likely contributors to motor symptoms in HE.

## Data Availability

All data produced in the present study are available upon reasonable request to the authors.

## Acknowledgment

The authors would like to thank Erika Rädisch (Department of Diagnostic and Interventional Radiology, University Hospital Düsseldorf) and Dr. Gerald Antoch (Department of Diagnostic and Interventional Radiology, University Hospital Düsseldorf) for support with the MR measurements. This study was supported by the Sonderforschungsbereich (SFB) 974 (TP B07) of the German Research foundation and by NIH grant R21 EB033516. The funding sources were not involved in the study design, collection, analysis, and interpretation of the presented data.

## CRediT authorship contribution statement

**Helge J. Zöllner**: Data acquisition, Formal Analysis, Investigation, Writing – Original Draft, Writing – Review & Editing, Visualization. **Thomas Thiel**: Data acquisition, Writing – Review & Editing, Visualization. **Markus S. Jördens**: Patient Recruitment, Clinical Evaluation, Formal Analysis, Writing – Review & Editing. **Sinyeob Ahn**: Data acquisition – Sequence Development, Writing – Review & Editing. **Dieter Häussinger**: Patient Recruitment, Formal Analysis, Writing – Review & Editing, Funding acquisition. **Markus Butz**: Data acquisition, Formal Analysis, Writing – Review & Editing, Project administration, Supervision, Funding acquisition. **Hans-Jörg Wittsack**: Conceptualization, Formal Analysis, Supervision, Writing – Review & Editing. **Alfons Schnitzler**: Formal Analysis, Project administration, Writing – Review & Editing, Supervision, Funding acquisition. **Georg Oeltzschner**: Conceptualization, Methodology, Writing – Review & Editing, Supervision, Funding acquisition.

