## Supplementary Material for "J-difference GABA-edited MRS reveals altered cerebello-thalamo-cortical metabolism in patients with hepatic encephalopathy"

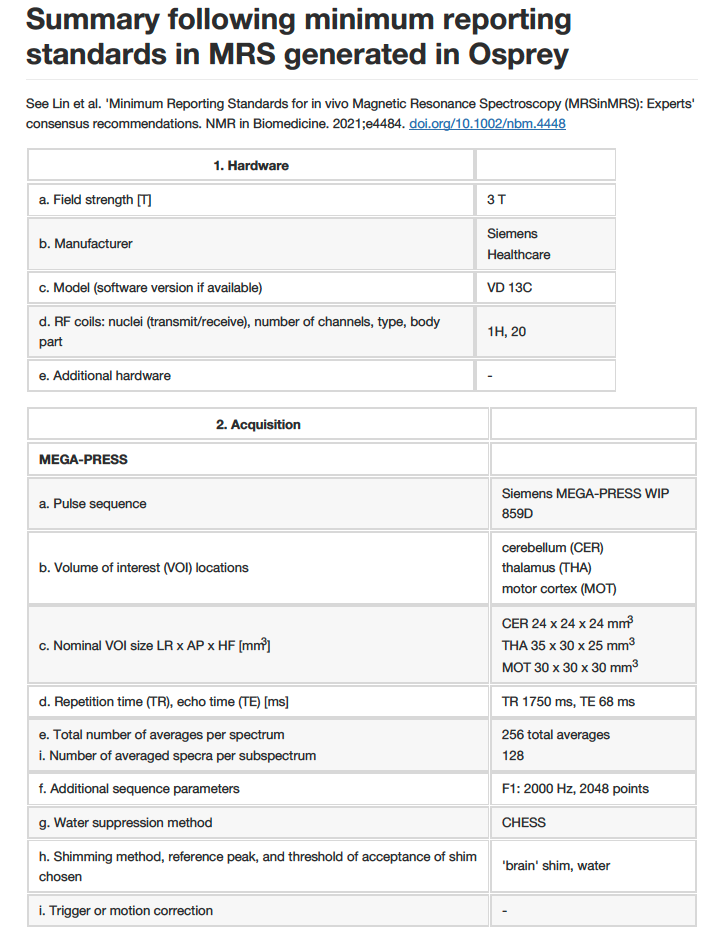

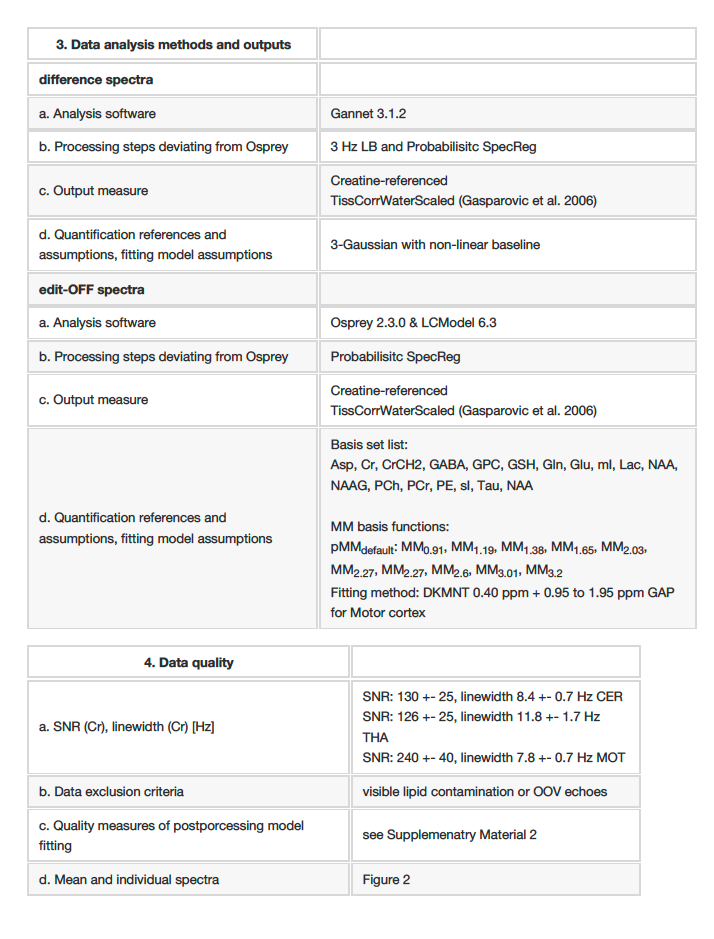

Supplementary Material 1 – MRSinMRS summary generated in Osprey

|  | **mean (SD)** | | | | | | | | | |
| --- | --- | --- | --- | --- | --- | --- | --- | --- | --- | --- |
| **CRLB (%)** | Gln | Glu | mI | GSH | tCho | tNAA | tCr | Asp | Tau | Scyllo |
| **Cerebellum** |  |  |  |  |  |  |  |  |  |  |
| *controls* | 25.1 (1.3) | 12.5 (2.4) | 10.4 (5.3) | 9.6 (3.7) | 5.1 (2.8) | 3.8 (0.9) | 3.0 (1.0) | 98 (125) | 74 (55) | 21.1 (7.7) |
| *mHE* | 21 (3) | 13 (2.5) | 11 (2.7) | 10.7 (4.0) | 6.0 (6.1) | 3.7 (1.2) | 2.7 (1.2) | 407 (519) | 216 (321) | 215 (320) |
| *HE* | 10.4 (3.6) | 11.2 (1.8) | 17.7 (11.7) | 7.9 (1.8) | 4.3 (2.5) | 3.0 (1.0) | 2.9 (1.1) | 29 (11) | 110 (267) | 180 (363) |
| **Thalamus** |  |  |  |  |  |  |  |  |  |  |
| *controls* | 18.8 (4.5) | 12.7 (3.1) | 9.9 (2.5) | 8.1 (2.2) | 3.7 (3.5) | 3.8 (1.2) | 2.9 (0.9) | 41 (38) | 494 (445) | 90 (213) |
| *mHE* | 13.7 (3.1) | 11.3 (4.0) | 10.7 (1.2) | 7.3 (1.2) | 4.0 (3.5) | 3.3 (0.6) | 3.0 (1.0) | 39 (37) | 36.7 (11) | 14.7 (6.5) |
| *HE* | 12.1 (5.9) | 11.5 (1.6) | 23.5 (25.8) | 7.0 (1.0) | 2.4 (0.9) | 3.4 (0.9) | 3.1 (1.2) | 29 (24) | 149 (272) | 346 (455) |
| **Motor cortex** |  |  |  |  |  |  |  |  |  |  |
| *controls* | 28.9 (10.9) | 6.8 (0.7) | 7.9 (1.2) | 6.9 (0.6) | 4.4 (0.9) | 2.7 (0.4) | 2.2 (0.4) | 11.7 (1.4) | 79 (29) | 15 (7.2) |
| *mHE* | 16 (1.4) | 7.5 (0.7) | 7.5 (0.7) | 7.0 (0) | 2.5 (0.7) | 3.0 (0) | 2.5 (0.7) | 10 (1.4) | 36 (9.2) | 14 (6.4) |
| *HE* | 14.8 (10.5) | 7.9 (1.5) | 96.8 (28) | 6.9 (1.2) | 4.3 (1.2) | 2.9 (1.2) | 2.6 (0.8) | 10.6 (2.2) | 59 (37) | 190 (377) |

Supplementary Material 2 – Metabolite Cramer-Rao Lower Bound (CRLB) of the linear-combination modeling estimates of the edit-OFF spectrum. Note inverse relation between high concentrations and lower CRLB, e.g. increased glutamine estimates in overt HE patients lead to lower CRLBs.

| **Creatine** | **mean (SD)** | | | | | | | | | |
| --- | --- | --- | --- | --- | --- | --- | --- | --- | --- | --- |
| **referenced** | Gln | Glu | mI | GSH | tCho | tNAA | tCr | Asp | Tau | Scyllo |
| **Cerebellum** |  |  |  |  |  |  |  |  |  |  |
| *controls* | .32 (.11) | .93 (.15) | .82 (.13) | .20 (.06) | .26 (.03) | 1.04 (.08) | - | .19 (.11) | .17 (.08) | .04 (.01) |
| *mHE* | .47 (.19) | 1.05 (.19) | .87 (.21) | .24 (.04) | .26 (.03) | 1.34 (.28) | - | .13 (.18) | .24 (.21) | .03 (.03) |
| *HE* | **.76 (.34)** | 1.00 (.19) | **.41 (.20)** | .22 (.05) | .22 (.05) | 1.09 (.14) | - | **.32 (.12)** | .23 (.10) | .02 (.02) |
| **Thalamus** |  |  |  |  |  |  |  |  |  |  |
| *controls* | .46 (.12) | 1.24 (.30) | .85 (.17) | .25 (.05) | .32 (.03) | 1.29 (.11) | - | .33 (.11) | .21 (.34) | .03 (.02) |
| *mHE* | .64 (.15) | 1.48 (.27) | .85 (.08) | .30 (.09) | .33 (.07) | 1.33 (.10) | - | .45 (.30) | .65 (.69) | .06 (.02) |
| *HE* | **.95 (.38)** | **1.70 (.43)** | **.56 (.22)** | **.31 (.05)** | **.28 (.07)** | 1.39 (0.19) | - | **.59 (.23)** | .44 (.29) | .02 (.02) |
| **Motor cortex** |  |  |  |  |  |  |  |  |  |  |
| *controls* | .19 (.05) | 1.00 (.05) | .86 (.08) | .22 (.02) | .26 (.03) | 1.55 (.09) | - | .65 (.08) | .07 (.03) | .04 (.01) |
| *mHE* | .36 (.05) | 1.01 (.09) | .82 (.01) | .24 (.03) | .27 (.03) | 1.61 (.19) | - | .93 (.13) | .18 (.05) | .05 (.03) |
| *HE* | **.72 (.53)** | 1.01 (.12) | **.44 (.26)** | .25 (.04) | **.24 (.04)** | 1.57 (.29) | - | **.90 (.23)** | .12 (.05) | **.02 (.01)** |

Supplementary Material 3 – Creatine-referenced metabolite estimates (mean (SD)) of the edit-OFF spectra quantified with linear-combination modeling. A HE-typical metabolite profile (increased Gln accompanied by mI reduction) was consistently found in all MRS voxels. Additionally, increased GSH (thalamus and motor cortex) and reduced tCho (cerebellum, thalamus & motor cortex) were found in some voxels. Further, increased aspartate levels were found in all three voxels and significantly decreased scyllo-inositol levels were found in the motor cortex of overt HE patients. Bold letters indicate significant (p_adj_ < .05) differences compared to healthy controls after multiple comparison corrections.

| **water** | **mean (SD)** | | | | | | | | | |
| --- | --- | --- | --- | --- | --- | --- | --- | --- | --- | --- |
| **referenced** | Gln | Glu | mI | GSH | tCho | tNAA | tCr | Asp | Tau | Scyllo |
| **Cerebellum** |  |  |  |  |  |  |  |  |  |  |
| *controls* | 6.4 (.2.2) | 18.7 (3.3) | 12.4 (3.0) | 4.6 (1.1) | 4.0 (.4) | 16.3 (1.3) | 19.4 (1.2) | 4.2 (2.3) | 4.0 (2.0) | .7 (.2) |
| *mHE* | 7.2 (.8) | 19.4 (2.5) | 11.8 (1.3) | 5.1 (1.2) | 3.9 (0.7) | 18.3 (2.4) | 19.3 (.02) | 4.2 (4.4) | 7.5 (3.3) | .6 (.8) |
| *HE* | **14.8 (6.4)** | 19.4 (3.9) | **6.3 (3.3)** | 4.8 (1.0) | **3.4 (.7)** | 16.2 (2.2) | **18.5 (1.3)** | **7.2 (2.8)** | 4.7 (2.4) | .4 (.3) |
| **Thalamus** |  |  |  |  |  |  |  |  |  |  |
| *controls* | 6.6 (1.6) | 17.5 (3.6) | 9.5 (2.1) | 4.0 (.8) | 3.5 (.2) | 14.3 (.9) | 13.7 (.9) | 5.0 (1.6) | 3.2 (5.2) | .5 (.3) |
| *mHE* | 8.7 (1.5) | 20.4 (3.7) | 9.1 (1.2) | 4.5 (.8) | 3.4 (.3) | 14.0 (1.2) | 13.3 (2.0) | 6.9 (5.1) | 8.8 (8.1) | .8 (.3) |
| *HE* | **13.0 (5.7)** | **23.2 (6.5)** | **5.9 (2.5)** | **4.8 (.8)** | **3.0 (.5)** | **13.1 (2.1)** | **12.6 (1.1)** | **8.2 (3.6)** | 6.2 (3.8) | .3 (.3) |
| **Motor cortex** |  |  |  |  |  |  |  |  |  |  |
| *controls* | 2.9 (.8) | 14.6 (.9) | 9.7 (1.0) | 3.6 (.2) | 2.9 (.2) | 17.5 (1.1) | 14.0 (.8) | 10.2 (1.4) | 1.25 (.51) | .6 (.1) |
| *mHE* | 5.2 (.3) | 14.4 (.2) | 9.1 (.6) | 3.8 (.1) | 3.0 (.6) | 17.4 (.6) | 13.7 (1.1) | 14.3 (3.1) | 2.8 (.6) | .7 (.3) |
| *HE* | **9.5 (7.6)** | **13.0 (1.2)** | **3.9 (2.2)** | 3.5 (.5) | **2.4 (.5)** | **15.4 (2.3)** | **12.2 (.9)** | 12.5 (3.6) | 1.49 (.59) | **.2 (.1)** |

Supplementary Material 4 – Water-referenced metabolite estimates (mean (SD)) of the edit-OFF spectra quantified with linear-combination modeling. A HE-typical metabolite profile (increased Gln accompanied by mI reduction) was consistently found in all MRS voxels. Additionally, increased GSH (thalamus and motor cortex) and reduced tCho (cerebellum, thalamus & motor cortex) were found in some voxels. Further, increased aspartate levels were found in all three voxels and significantly decreased scyllo-inositol levels were found in the motor cortex of overt HE patients. Bold letters indicate significant (p_adj_ < .05) differences compared to healthy controls after multiple comparison corrections.

| c**reatine** | **weighted partial correlation r** | | | | | | | | | |
| --- | --- | --- | --- | --- | --- | --- | --- | --- | --- | --- |
| **referenced** | Gln | Glu | mI | GSH | tCho | tNAA | tCr | Asp | Tau | Scyllo |
| **Cerebellum** |  |  |  |  |  |  |  |  |  |  |
| *CFF* | **-0.54** | -0.13 | **0.65** | -0.26 | 0.32 | -0.22 | - | **-0.68** | -0.15 | 0.18 |
| *tPEG score* | **0.63** | 0.25 | **-0.75** | 0.35 | -0.33 | 0.16 | - | 0.31 | 0.05 | -0.15 |
| *NH3* | 0.61 | -0.13 | -0.62 | 0.42 | -0.27 | -0.75 | - | -0.24 | -0.31 | 0.07 |
| **Thalamus** |  |  |  |  |  |  |  |  |  |  |
| *CFF* | **-0.56** | **-0.52** | **0.50** | -0.45 | 0.26 | -0.36 | - | **-0.50** | -0.40 | 0.00 |
| *tPEG score* | **0.76** | **0.61** | -0.42 | **0.47** | -0.31 | 0.25 | - | **0.50** | -0.01 | -0.18 |
| *NH3* | 0.57 | -0.36 | -0.19 | -0.17 | -0.33 | -0.39 | - | -0.01 | -0.26 | 0.23 |
| **Motor cortex** |  |  |  |  |  |  |  |  |  |  |
| *CFF* | -0.50 | 0.11 | 0.77 | -0.36 | 0.18 | -0.10 | - | **-0.63** | -0.30 | **0.50** |
| *tPEG score* | 0.74 | -0.19 | -0.80 | 0.35 | -0.47 | -0.05 | - | 0.48 | 0.37 | **-0.69** |
| *NH3* | 0.62 | -0.38 | -0.10 | 0.02 | -0.27 | -0.46 | - | 0.11 | 0.18 | 0.00 |
| ***water concentrations*** | Gln | Glu | mI | GSH | tCho | tNAA | tCr | Asp | Tau | Scyllo |
| **Cerebellum** |  |  |  |  |  |  |  |  |  |  |
| *CFF* | **-0.49** | -0.02 | **0.55** | -0.12 | 0.41 | -0.08 | 0.19 | **-0.63** | -0.19 | 0.40 |
| *tPEG score* | **0.58** | 0 | **-0.70** | 0.12 | **-0.49** | -0.19 | **-0.54** | 0.23 | 0.07 | -0.43 |
| *NH3* | 0.40 | -0.20 | -0.39 | 0.33 | -0.31 | -0.65 | -0.48 | -0.32 | -0.32 | 0.14 |
| **Thalamus** |  |  |  |  |  |  |  |  |  |  |
| *CFF* | **-0.52** | **-0.47** | **0.50** | -0.39 | 0.40 | -0.14 | 0.24 | **-0.45** | -0.38 | 0.06 |
| *tPEG score* | **0.74** | **0.53** | **-0.48** | 0.39 | **-0.57** | 0.03 | -0.23 | **0.49** | -0.05 | -0.23 |
| *NH3* | 0.48 | -0.41 | -0.26 | -0.32 | -0.71 | **-0.60** | -0.16 | -0.06 | -0.26 | 0.29 |
| **Motor cortex** |  |  |  |  |  |  |  |  |  |  |
| *CFF* | -0.46 | **0.66** | **0.80** | 0.18 | 0.49 | 0.42 | **0.69** | -0.43 | -0.18 | **0.57** |
| *tPEG score* | **0.74** | -0.62 | **-0.80** | -0.02 | **-0.62** | -0.44 | -0.44 | 0.35 | 0.29 | **-0.73** |
| *NH3* | 0.63 | -0.26 | 0 | 0.16 | -0.30 | -0.36 | -0.02 | 0.19 | 0.23 | 0.07 |

Supplementary Material 5 – Correlation analysis between metabolite estimates (creatine- and water referenced) and clinical metrics. Weighted partial correlations were used to account for participant’s age as a covariate and model uncertainty (CRLB) as weight.
